# Prevalence and determinants of cigarette smoking and smoking frequency among people with tuberculosis in Lesotho: evidence from a nationwide cross-sectional survey

**DOI:** 10.1101/2025.10.07.25337531

**Authors:** Calista O. Dozie-Nwakile, Alphonsus O. Ogbuabor, Nwanneka C. Ghasi, Daniel C. Ogbuabor

**Affiliations:** Department of Medical Laboratory Sciences, Faculty of Health Sciences and Technology, University of Nigeria, Enugu Campus, Enugu, Nigeria; Department of Medical Laboratory Sciences, Faculty of Allied Health Sciences, College of Medicine, Enugu State University of Science and Technology, Enugu, Nigeria; Department of Management, Faculty of Business Administration, University of Nigeria, Enugu Campus, Enugu, Enugu, Nigeria; Department of Health Administration and Management, Faculty of Health Sciences and Technology, University of Nigeria, Enugu Campus, Enugu, Enugu, Nigeria

**Author notes:** Corresponding author: Alphonsus C. Ogbuabor, Department of Medical Laboratory Sciences, Faculty of Allied Health Sciences, College of Medicine, Enugu State University of Science and Technology, Enugu, Nigeria. Country of authors: Nigeria, Email addresses: COD, NCG, DCO. **Declarations**. **Ethics approval and consent to participate** Since this study was a secondary analysis of the Lesotho Demographic and Health surveys (LDHS) data, which are publicly available, the study did not require any ethical approval. Only DHS program authorization was requested to download the dataset. **Consent for publication** Not applicable. **Availability of data and material** The data used for this study are from the Lesotho Demographic and Health surveys (LDHS) 2023-24 and are publicly available here: https://dhsprogram.com/data/available-datasets.cfm Data was accessed by the researchers upon registration. **Competing interests** The authors report no conflict of interest. **Funding** The authors received no external funding for this study. **Authors’ contributions** COD, AOO, CNG, and DCO contributed to the conceptualisation of the study. COD, AOO, NCG, and DCO conducted the literature review. AOO, NCG and DCO abstracted and analysed the data. COD, AOO and DCO summarized the data and drafted the manuscript. COD, AOO, NCG, and DCO critically reviewed, discussed, and modified the intellectual content of the article. All authors read and approved the final manuscript. **Acknowledgements** We thank the DHS Program for availing us access to the data.

## Abstract

**Background:** Low-and middle-income countries face a significant dual burden of tuberculosis (TB) and cigarette smoking. Nevertheless, studies examining the prevalence and determinants of smoking status and frequency among people with TB (PWTB) are scarce in sub-Saharan Africa, including Lesotho. Therefore, this study evaluated the prevalence and determinants of smoking status and frequency among people with TB in Lesotho.

**Methods:** Data on people with TB from the 2023 Lesotho Demographic and Health Survey were used in this study (n = 224). The data were adjusted for sampling weight, stratification, and cluster sampling design. The outcome variables were smoking status and smoking frequency (daily smoking and occasional smoking). The predictor variables included socio-demographic factors, household characteristics, alcohol use, TB-related stigma, and mental health status. We evaluated the association between outcome and predictor variables using Pearson’s chi-squared test and complex sample logistics regression. Statistical significance was set at a p-value < 0.05.

**Results:** The prevalence of cigarette smoking among PWTB is 32.2%. The prevalence of daily and occasional smoking among PWTB is 24.9% and 7.3%, respectively. Being male (AOR=120.15, 95%CI:30.53-472.81, p<0.001), no media exposure (AOR=21.96, 95%CI:5.79-83.31, p<0.001), low media exposure (AOR=4.76, 95%CI:1.33-17.09, p=0.017), alcohol use (AOR=12.64, 95%CI:3.24-49.35, p<0.001), and mild depression (AOR=3.28, 95%CI: 3.28, 95%CI: 1.04-10.38, p=0.043) increased the odds of smoking among PWTB. Nevertheless, moderate/severe depression (AOR=0.06, 95%CI: 0.00-0.73, p=0.027) reduced the likelihood of smoking among PWTB. Being male (AOR=234.07, 95%CI:108.69-504.08, p<0.001), no media exposure (AOR=18.93, 95%CI:4.58-78.28, p<0.001), and alcohol use (AOR=21.26, 95%CI:3.79-119.23, p=0.001) increased the odds of smoking among PWTB. In contrast, moderate/severe depression (AOR=0.00, 95%CI: 0.00-0.00, p<0.001) reduced the likelihood of smoking among PWTB. Being male (AOR=46.55, 95%CI:4.69-461.61, p=0.001), no media exposure (AOR=94.66, 95%CI:7.60-1179.67, p=0.001), and low media exposure (AOR=26.34, 95%CI:2.54-272.98, p=0.007), and mild depression (AOR=6.18, 95%CI:1.43-26.66, p=0.015) increased the odds of smoking among PWTB.

**Conclusion:** The high prevalence of smoking among PWTB needs to improve in Lesotho. Policies and programs to reduce smoking among PWTB must target the male gender, improve media exposure to anti-tobacco campaigns, reduce alcohol consumption and improve mental health among people with TB.

## Background

Low-and middle-income countries (LMICs) face a significant dual burden of tuberculosis (TB) and cigarette smoking[1]. In 2023, approximately 10.8 million people globally were affected by TB, leading to an estimated 1.25 million deaths[2]. Despite a decline in incidence rates, sub-Saharan Africa (SSA) continues to host over 50% of the countries with a high burden of TB, including Lesotho[2]. Although there has been a 15% reduction in TB incidence, Lesotho’s rate remains high at 664 cases per 100,000 people[2]. Equally, more than 80% of the 1.3 billion smokers worldwide live in low-and middle-income countries, contributing to over eight million deaths each year from tobacco-related causes[3]. In SSA, the prevalence of cigarette smoking stands at 7.1%, which is significantly lower than Lesotho’s 18.3%[3]. Smoking accounts for more than 20% of global TB incidence and about 1,400 TB cases in Lesotho[2]. Current smokers are more likely to have both symptomatic and subclinical TB[4]. Ever-smokers, current smokers, and past smokers have an elevated risk of TB in comparison to never-smokers[5]. Consequently, the End TB strategy emphasizes the importance of TB screening for smokers and incorporates smoking cessation into standard TB care practices[6].

Studies conducted in various settings have shown that tobacco smoking among people with tuberculosis (TB) is significantly associated with several adverse health outcomes. These include longer diagnostic delays[7], a higher likelihood of positive sputum smears and cavitary TB[7–10], prolonged sputum conversion time[11, 12], increased rates of TB relapse or recurrence[1, 13–15], greater chances of drug-resistant TB, poor adherence to treatment[16, 17], and higher rates of loss to follow-up[16–19], treatment failure[13, 14, 16–19], and death[1, 13, 14, 16, 18, 19]. In high-burden TB countries, smoking contributes to 15% of TB-related mortality[20]. Furthermore, a systematic review revealed that smoking increases the likelihood of poor treatment outcomes by 51% globally and by 74% in LMICs[21]. However, a West African study found no significant differences in disease severity at the time of diagnosis and no notable differences in treatment outcomes, which are attributable to confounding socioeconomic factors[22].

The prevalence of smoking among TB patients varies widely. In Iran, the prevalence is 14%[23]. In Pakistan, the prevalence ranges from 19.8% in pulmonary TB patients to 22.0% in all TB patients[24]. While a prevalence of 21.8% was found in India[18], Bangladesh’s ranges from 24.4% in pulmonary TB patients to 28.3% in all TB patients[24]. Some studies have indicated even higher rates, exceeding 30%, in Brazil, Jordan, Spain, and Georgia[10, 17, 19, 25]. In SSA, the prevalence of smoking among people with TB ranges from 15.2% to 26% in Uganda[26, 27], whereas South Africa exhibits alarmingly high rates between 56% and 82%[28–30]. Additional studies indicate prevalence rates of 16.2% in Ethiopia[31], 25% in Botswana[32], and 30% in Gabon[7]. These findings highlight a need for targeted smoking cessation interventions for individuals diagnosed with TB in different settings. A comprehensive understanding of the social determinants of smoking among people with TB is essential for developing and implementing focused smoking cessation interventions for those most at risk. Existing literature suggests that age[33], male gender[10, 16, 25, 30, 32–34], urban residency[16], lower education[30], employment status[16], alcohol use[10, 16, 34, 35], depression[32], and financial difficulties[25] are risk factors for smoking among people with TB.

There is a paucity of studies examining the prevalence and determinants of smoking among people with TB globally, particularly in SSA. Most African studies utilize facility-based surveys that are not nationally representative or do not focus on social determinants[7, 12, 22, 26, 27, 29–31, 33, 34]. In Lesotho, existing health system research on TB focus on evaluating determinants of treatment outcome[36, 37], understanding the barriers to TB care[38], and knowledge and attitude towards TB[39]. Lesotho’s National Tuberculosis Strategic Plan 2018-2022 did not prioritize smoking as a critical social determinant of TB[40]. To our knowledge, no published study has assessed the prevalence and predictors of smoking among people with TB in Lesotho despite having the highest incidence of TB in SSA[2], and a high prevalence of smoking in the general population[3]. Therefore, our study fills an important gap in the literature by presenting new evidence regarding the prevalence and determinants of smoking among people with TB in Lesotho utilizing a nationally representative population-based survey. Such evidence would be helpful to TB policymakers, program managers, and service providers in prioritizing smoking in TB strategic plans and integrating smoking cessation interventions into routine TB care and prevention strategies.

## Methods

### Study setting

Lesotho is a small lower-middle-income country with a population of 2.3 million and a gross domestic product (GDP) per capita of USD1,300. Almost two-thirds (67%) of the population lives in rural areas, and females account for 51% of the population. There are high levels of inequality in Lesotho, with a Gini coefficient of 44.6%, and 24% of people live in extreme poverty[41]. Lesotho has ten administrative districts. Each district is subdivided into constituencies, and each constituency into community councils. The districts serve as TB programme coordinating and reporting units under the leadership of the ten District Health Management Teams. The country achieved the 2020 milestone of the End TB strategy (a 20% reduction compared with the 2015 baseline). Nevertheless, the TB programme has a 68% case identification gap and a 76% treatment success rate.

### Study design

We analysed secondary data from the 2023-24 Lesotho Demographic and Health Survey (LDHS). The primary study used a cross-sectional, household survey design. We accessed the LDHS data set upon request through the ICF Macro DHS data management portal.

### Sampling strategy

The primary study calculated the sample size of households (n = 10,000) using information obtained from the 2014 LDHS. The sample calculations used information obtained from the 2014 LDHS: the average number of women age 15–49 per household was 0.816 in urban areas and 0.687 in rural areas, the average number of men age 15–59 per household was 0.683 in urban areas and 0.663 in rural areas, the household completion rate was 94.6%, the response rate among women age 15–49 was 97.1%, and the response rate among menage 15–59 was 94%.

The 2023-24 LDHS used a two-stage stratified sampling technique to select the households. The sampling frame consisted of households listed in Lesotho’s 2016 Population and Housing Census (PHC) conducted by the Lesotho Bureau of Statistics (BoS). The primary sampling unit (PSU) is a cluster of enumeration areas. An enumeration area (EA) is a geographical area, usually a city block in urban areas or a village in rural areas, with an adequate number of households. Each district was stratified into urban and rural areas. In the first stage, the study selected 400 (176 urban and 224 rural) EAs from the sampling strata with probability proportional to EA size. The second stage’s sampling frame is the household list in each EA. In the second stage, 25 households were selected from every EA through equal probability systematic sampling, resulting in a total sample size of about 10,000 households (4,400 urban and 5,600 rural households). However, the study selected 9,976 households for the Women’s Questionnaire and 4,993 (from a subsample of half of the women’s households) for the Men’s Questionnaire. No replacements and no changes to the preselected households were allowed in the implementation stages to prevent bias.

### Data collection

The data collection took place over 3 months from 27 November 2023 to 29 February 2024 across the 10 districts of Lesotho. All women age 15–49 (n = 6,536) and all men age 15–59 (n = 3,304) in the subsample who were usual residents of the sampled households or stayed in the households on the night before the interview were eligible for interviews. Data were successfully collected from 6,413 women and 3,214 men, representing 98.1% and 97.3% of eligible women and men, respectively. The survey data were collected using tablet computers running the Android operating system and Census and Survey Processing System (CSPro) software. The data collectors comprised 15 field teams, each consisting of one team supervisor, three or four female interviewers, and one to three male interviewers. Each interviewer entered the answers to the survey questions into the tablets. Supervisors downloaded interview data to their tablet, checked it for completeness, and monitored fieldwork progress.

### Variables Outcome variable

The outcome variables were smoking status and smoking frequency at the time of the survey. In this study, smoking cigarettes included smoking combustible cigarettes such as manufactured cigarettes and hand-rolled cigarettes[42], but excluded cigars, cheroots, cigarillos, hookah, kreteks, and e-cigarettes. The study assessed current cigarette smoking using the specific question “Do you currently smoke cigarettes?” Respondents who answered “yes” to the question were classified as current cigarette smokers, while those who answered “no” were classified as women who do not currently smoke. Furthermore, the smokers were asked whether they smoked every day or only on some days. We recoded the smoking frequency into a binary variable: daily (every day) and occasional (some days) smoking.

### Predictor variables

We selected the predictor variables based on previous studies on smoking in Africa and the data availability in the DHS database. The factors included in this study are gender ( male and female), age (15-19, 20-29, 30-39, and ≥40 years), marital status (Never in a union, married/living with a partner, and divorced/separated/widowed), district, place of residence (urban and rural), highest education (no education, primary, secondary, and higher), religion (Catholic, Protestants, Pentecostal/other Christians, and Others), Ethnicity (Basotho or other), sex of household head (female and male), household size (≤5 and >5), media exposure (No, low, and high), ownership of mobile phone (yes, no), wealth index (poorest, poor, moderate, rich, richest), occupation (not working, professional/technical/managerial, clerical/sales/services, agricultural, and manual), employment (unemployed and employed), total children (no child, one, 2-4, and >4), health insurance ownership (No and Yes), alcohol use (No and Yes), wife beating (No and yes), and TB-related stigma (No stigma and Yes). Wife-beating attitude was measured using five variables describing respondents’ attitudes towards domestic violence, including whether beating was justified if the wife goes out without telling her husband, neglects the children, argues with her husband, refuses sex, and burns food[43]. People who answered’Yes’ and’Don’t know’ were coded as 1, while those who responded’No’ were coded as 0. Respondents were categorised into good gender attitude if they answered’No’ to all five variables, and poor gender attitude if they answered’Yes’ or’Don’t know’ to any of the five questions.

We derived a composite measure of TB-related stigma using three specific questions: whether a respondent believes TB is curable, keeps secret if a family member has TB, and is willing to work with someone with TB. Individuals who answered “yes” to questions regarding tuberculosis (TB) treatment options or their willingness to work with someone with TB were coded as’1’. In contrast, those who answered “no” or “don’t know/not sure/depends” were recoded as’0’. For the question regarding keeping TB a secret, respondents who answered “no” or “don’t know/not sure/depends” were recoded as’1’, while those who answered “yes” were recoded as’0’. We summed the scores after completing the recoding process. We classified individuals who obtained a total score of 3 as having “no stigma,” and those with scores below three as having “stigma.”

The 2023-24 LDHS screened respondents’ mental health using the Patient Health Questionnaire, or PHQ-9 (Kroenke & Spitzer, 2002). The sum of the scores on each of the nine items forms the PHQ-9 score. Each symptom in the PHQ-9 is assigned a score of 0, 1, 2, or 3 depending on how frequently the respondent reported experiencing the symptom in the 2 weeks preceding the survey: 0 – Never, 1 – Rarely, 2 – Often, and 3 – Always. PHQ-9 scores range from a minimum of 0 to a maximum of 27. Higher scores reflect more severe symptoms of depression. A PHQ score of 0–4 indicates minimal symptoms or no symptoms, while a score of 5–9 is considered mild, 10–14 is moderate, 15–19 is considered moderately severe, and 20–27 is considered severe. In our analysis, we recoded people with minimal or no symptoms to’0’, mild to’1’, moderate to moderately severe, and severe to’2’.

### Statistical analysis

We analysed the data using Statistical Package for Social Sciences (SPSS) version 20 (IBM Corp., Armonk, NY). Before analysis, we adjusted the data for sampling weights, stratification, and multistage sampling to account for the variation in sample allocation to the districts and provide representative population estimates. Individual sample weights were generated by dividing (v005 and mv005) by 1,000,000 before analysis to approximate the number of cases. We checked for the multicollinearity of the independent variables using variance inflation factors (VIF), and they ranged from 1.17 to 2.08. We reported descriptive statistics using frequencies, population estimates and percentages (weighted). We assessed the association between the outcome variables (smoking status and frequency) and the independent variables using the Chi-square test. All variables whose univariable test had a p-value of less than 0.05 were included as covariates in the multivariable complex sample logistic regression model to determine the adjusted effect of each predictor variable on smoking status or frequency. The results of regression analysis were presented by crude/unadjusted odds ratio (COR) and adjusted odds ratio (AOR) with 95% confidence intervals (CIs), and p-values of less than 0.05 were considered significant. We tested the model fit using McFadden’s R-squared because it compares the likelihood-ratio of the current model to a model without any covariates and represents the amount of variation explained by the current model[44]. The McFadden test statistic for smoking status and smoking frequency was 0.55 and 0.47, respectively, indicating a good model fit[44].

### Ethical consideration

We did not obtain further ethical approval as this was a secondary data analysis. The 2023–24 LDHS protocol received clearance from the ICF Institutional Review Board ethics committee and the Lesotho Ministry of Health Research and Ethics Committee. Additionally, interviewers obtained informed consent from participants before conducting interviews.

## Results

### Basic characteristics of people with TB

Table 1 shows the basic characteristics of the respondents. Over 60% of the respondents were females, married or living with a partner, and employed. About 77% of the respondents lived in three of the country’s nine districts (Maseru, Leribe, and Berea districts). Most respondents are Christians, have at least basic education, hail from Basotho tribe, and own a mobile phone. Most household have five or less people. Almost 35% of households are poor. Health insurance coverage is low. Approximately two-third of the respondents use alcohol.

**Table 1.** Basic characteristics of people with TBin Lesotho, 2023-24 (N=224)

| Basic characteristics |  | Frequency (n) | Percent (%) |
| --- | --- | --- | --- |
| Gender | Male | 123 | 54.9 |
|  | Female | 101 | 45.1 |
| Age group | 15-19 | 8 | 3.6 |
|  | 20-29 | 22 | 9.7 |
|  | 30-39 | 59 | 26.4 |
|  | 40 and above | 135 | 60.4 |
| Current marital status | Never in union | 37 | 16.4 |
|  | Currently in union/living with a woman | 144 | 64.2 |
|  | Formerly in union/living with a woman | 43 | 19.4 |
| District | Butha-Buthe | 7 | 3.3 |
|  | Leribe | 42 | 18.9 |
|  | Berea | 39 | 17.5 |
|  | Maseru | 93 | 41.6 |
|  | Mafeteng | 12 | 5.5 |
|  | Mohale's Hoek | 11 | 4.7 |
|  | Quthing | 5 | 2.4 |
|  | Qacha's Nek | 4 | 1.7 |
|  | Mokhotlong | 2 | 0.8 |
|  | Thaba-Tseka | 8 | 3.5 |
| Type of place of residence | Urban | 118 | 52.7 |
|  | Rural | 106 | 47.3 |
| Educational level | No education | 15 | 6.9 |
|  | Primary | 90 | 40.0 |
|  | Secondary | 104 | 46.5 |
|  | Higher | 15 | 6.6 |
| Religion | Roman Catholic | 94 | 41.9 |
|  | Protestant | 64 | 28.5 |
|  | Pentecostal/other Christians | 54 | 24.0 |
|  | Other religions | 12 | 5.5 |
| Ethnicity | Basotho | 215 | 95.8 |
|  | Others | 9 | 4.2 |
| Sex of household head | Male | 170 | 75.8 |
|  | Female | 54 | 24.2 |
| Household size | 5 or <5 | 163 | 72.7 |
|  | >5 | 61 | 27.3 |
| Media exposure | No media exposure | 32 | 14.2 |
|  | Low media exposure | 117 | 52.4 |
|  | High media exposure | 75 | 33.5 |
| Owns a mobile telephone | No | 47 | 20.8 |
|  | Yes | 178 | 79.2 |
| Wealth index for urban/rural | Poorest | 39 | 17.4 |
|  | Poorer | 43 | 19.2 |
|  | Middle | 53 | 23.6 |
|  | Richer | 31 | 14.0 |
|  | Richest | 58 | 25.8 |
| Occupation | Not working | 42 | 18.9 |
|  | Professional/Technical/Managerial | 7 | 3.3 |
|  | Clerical/Sales/Services | 42 | 18.7 |
|  | Agricultural workers | 21 | 9.2 |
|  | Manual workers | 112 | 49.9 |
| Currently employed | No | 73 | 32.8 |
|  | Yes | 151 | 67.2 |
| Total Children | No child | 39 | 17.2 |
|  | One child | 52 | 23.0 |
|  | 2-4 children | 103 | 46.1 |
|  | >4 children | 31 | 13.6 |
| Covered by health insurance | No | 183 | 81.6 |
|  | Yes | 41 | 18.4 |
| Alcohol use | Never/No | 64 | 28.8 |
|  | Yes | 160 | 71.2 |
| Wife beating | No | 174 | 77.5 |
|  | Yes | 51 | 22.5 |
| TB-related Stigma | No stigma | 193 | 86.0 |
|  | Stigma | 31 | 14.0 |
| Mental health status | No depression | 160 | 71.2 |
|  | Mild | 51 | 23.0 |
|  | Moderate/severe | 13 | 5.9 |

### Prevalence of smoking among people with TB

The prevalence of cigarette smoking among people with TB is 32.2% (Table 2). Smoking prevalence among people with TB differed significantly by sex of household head, media exposure, occupation, alcohol use and attitude to wife beating.

**Table 2.**
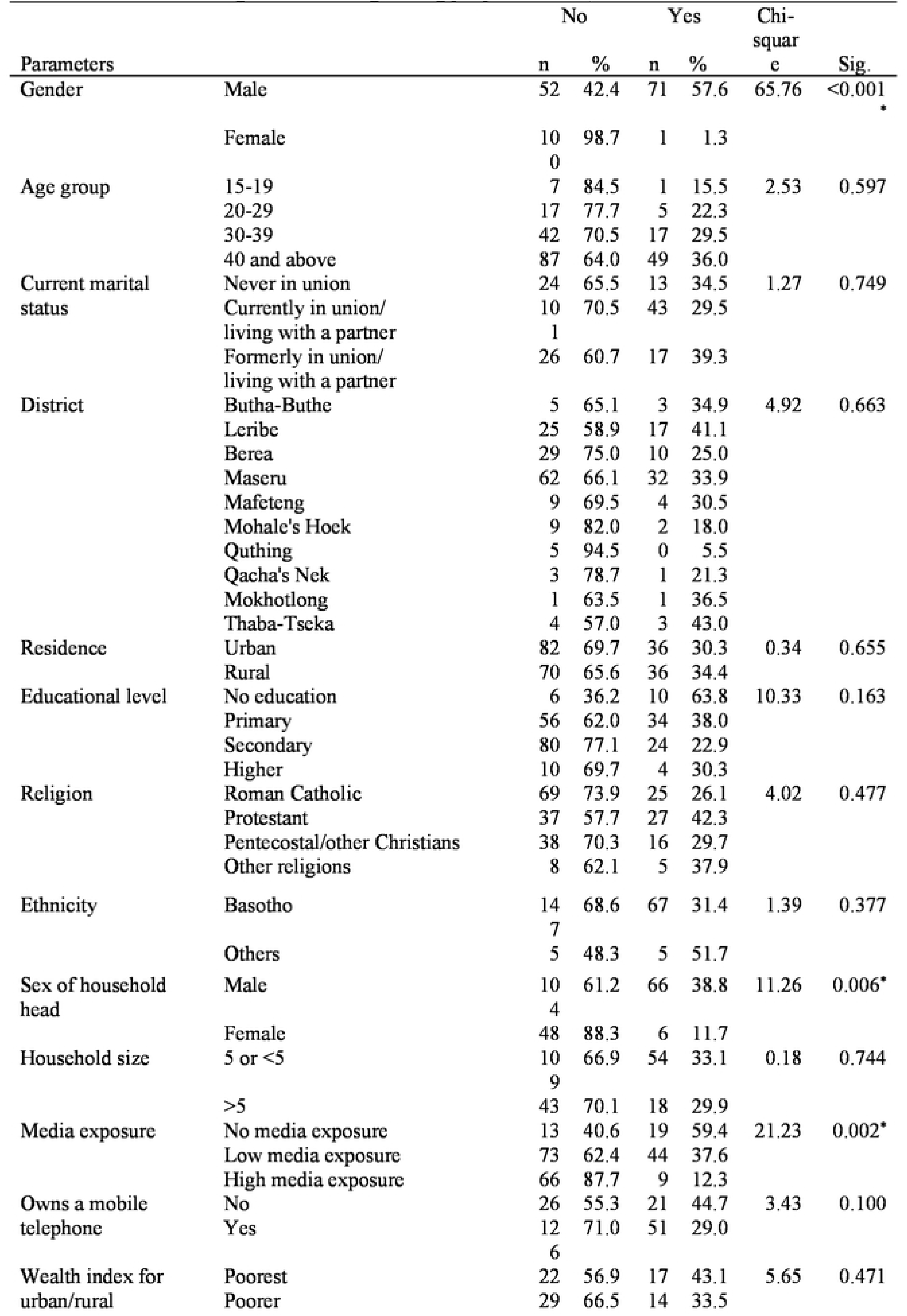
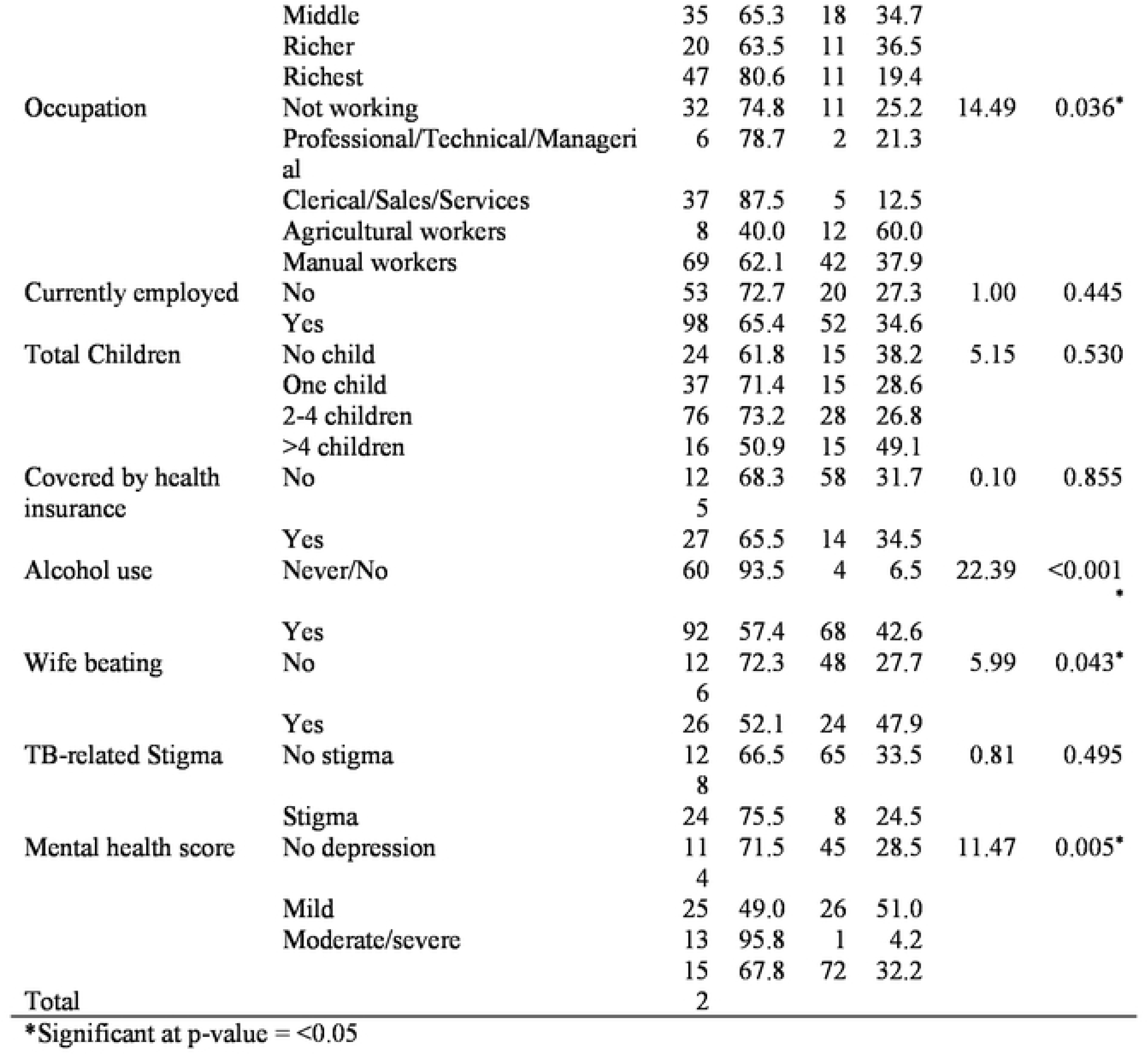
Prevalence of cigarette smoking among 11eo11Ie with TB, 2023-24.

### Prevalence of smoking frequency among people with TB

The prevalence of daily and occasional smoking among people with TB is 24.9% and 7.3%, respectively (Table 3). The prevalence of smoking frequency among people with TB differed significantly by sex of household head, media exposure, alcohol use and attitude to wife beating.

**Table 3.**
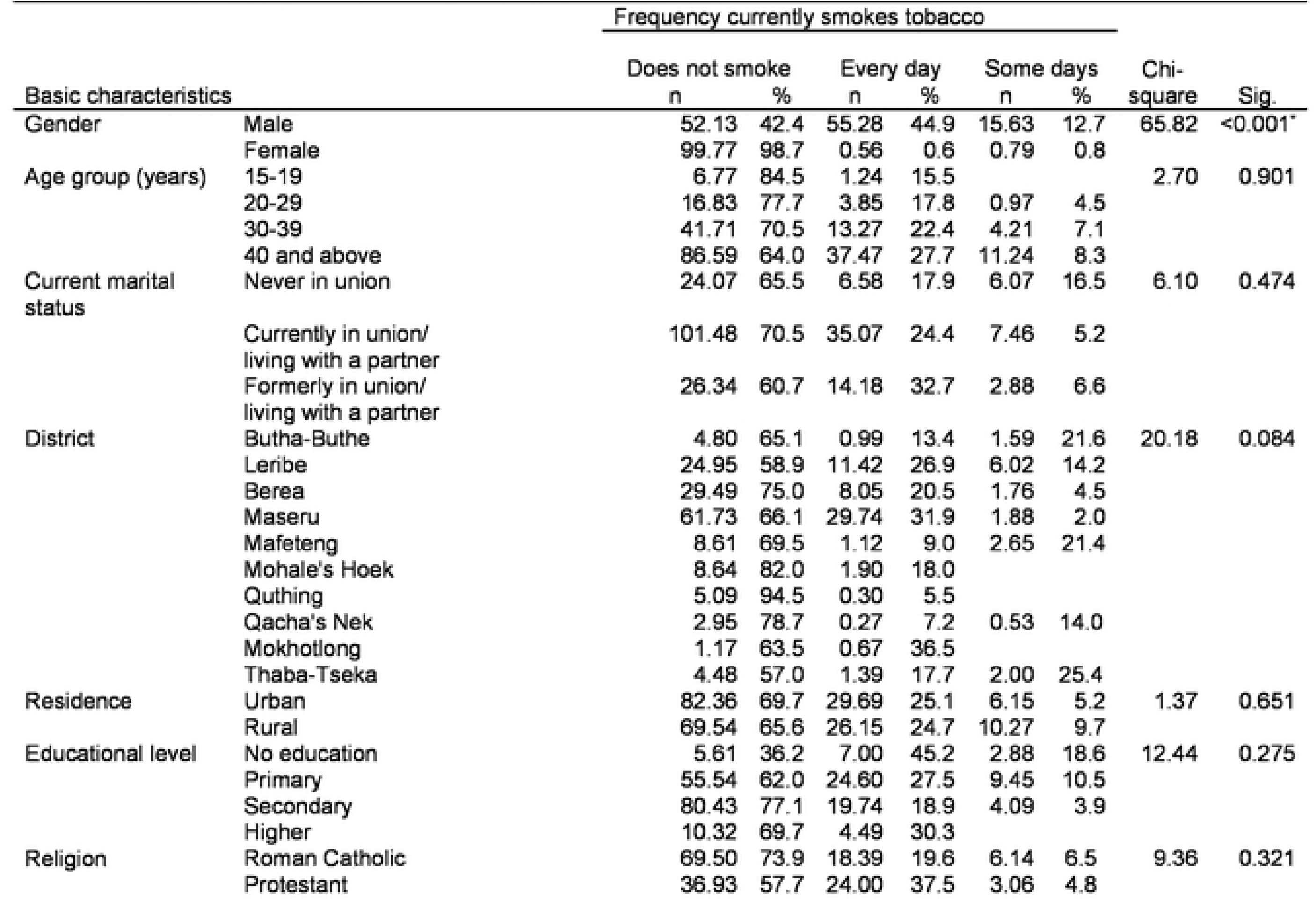
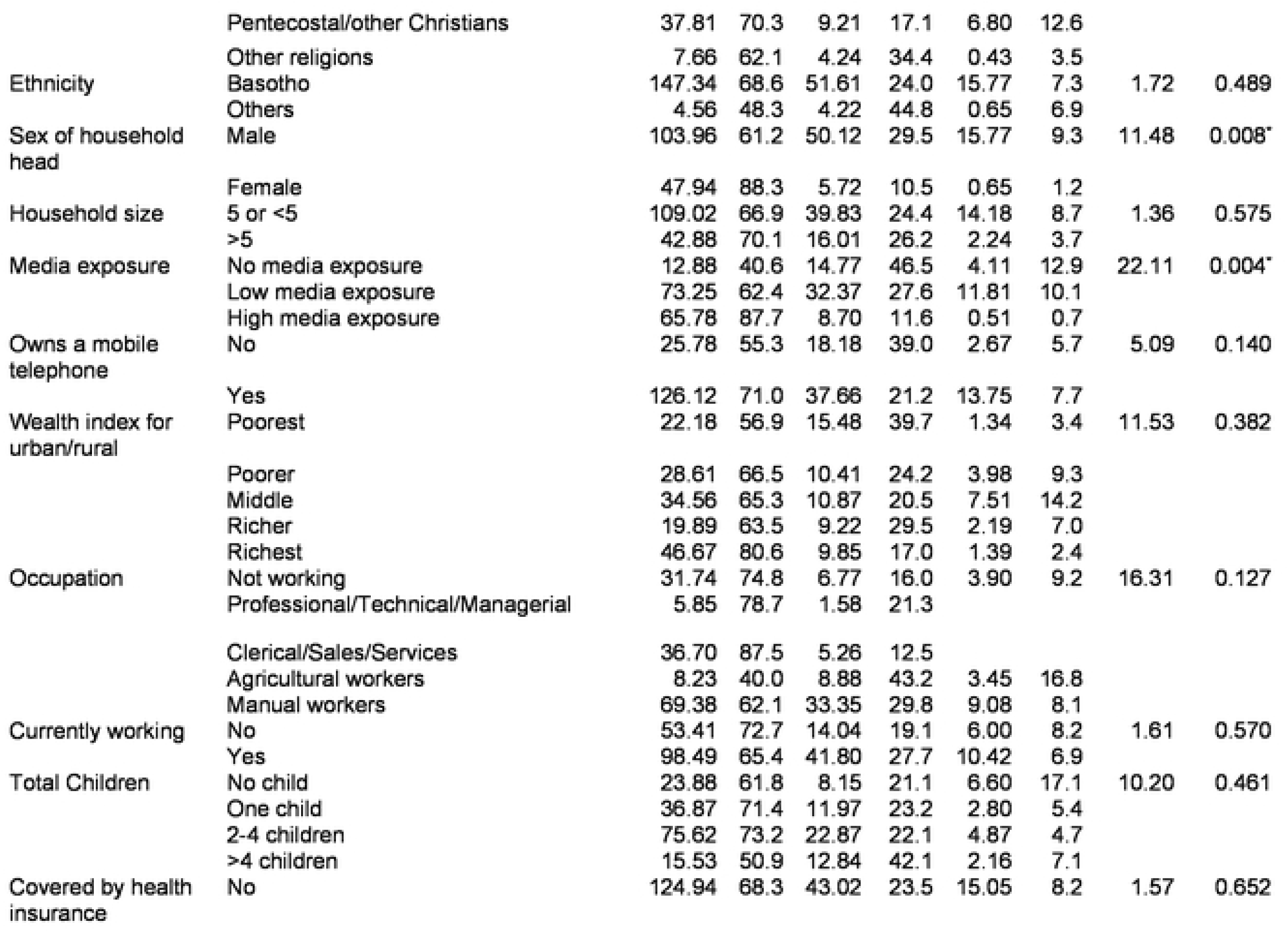
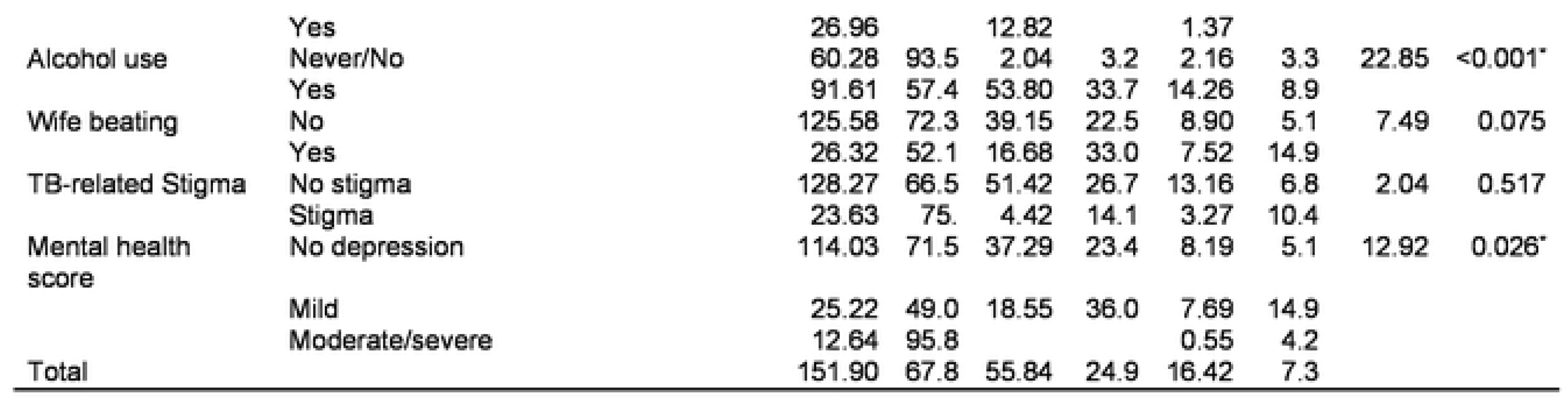
Prevalence of ci9arette smokin9 freguencl amons <u>peoele</u> with TB,2023-24.

|  |  | Frequency currently smokes tobacco |  |  |  |  |  |  |  |
| --- | --- | --- | --- | --- | --- | --- | --- | --- | --- |
| Basic characteristics |  | Does not smoke |  | Every day |  | Some days |  | Chi-square | Sig. |
|  |  | n | % | n | % | n | % |  |  |
| Gender | Male | 52.13 | 42.4 | 55.28 | 44.9 | 15.63 | 12.7 | 65.82 | <0.001* |
|  | Female | 99.77 | 98.7 | 0.56 | 0.6 | 0.79 | 0.8 |  |  |
| Age group (years) | 15-19 | 6.77 | 84.5 | 1.24 | 15.5 |  |  | 2.70 | 0.901 |
|  | 20-29 | 16.83 | 77.7 | 3.85 | 17.8 | 0.97 | 4.5 |  |  |
|  | 30-39 | 41.71 | 70.5 | 13.27 | 22.4 | 4.21 | 7.1 |  |  |
|  | 40 and above | 86.59 | 64.0 | 37.47 | 27.7 | 11.24 | 8.3 |  |  |
| Current marital status | Never in union | 24.07 | 65.5 | 6.58 | 17.9 | 6.07 | 16.5 | 6.10 | 0.474 |
|  | Currently in union/<br>living with a partner | 101.48 | 70.5 | 35.07 | 24.4 | 7.46 | 5.2 |  |  |
|  | Formerly in union/<br>living with a partner | 26.34 | 60.7 | 14.18 | 32.7 | 2.88 | 6.6 |  |  |
| District | Butha-Buthe | 4.80 | 65.1 | 0.99 | 13.4 | 1.59 | 21.6 | 20.18 | 0.084 |
|  | Leribe | 24.95 | 58.9 | 11.42 | 26.9 | 6.02 | 14.2 |  |  |
|  | Berea | 29.49 | 75.0 | 8.05 | 20.5 | 1.76 | 4.5 |  |  |
|  | Maseru | 61.73 | 66.1 | 29.74 | 31.9 | 1.88 | 2.0 |  |  |
|  | Mafeteng | 8.61 | 69.5 | 1.12 | 9.0 | 2.65 | 21.4 |  |  |
|  | Mohale's Hoek | 8.64 | 82.0 | 1.90 | 18.0 |  |  |  |  |
|  | Quthing | 5.09 | 94.5 | 0.30 | 5.5 |  |  |  |  |
|  | Qacha's Nek | 2.95 | 78.7 | 0.27 | 7.2 | 0.53 | 14.0 |  |  |
|  | Mokhotlong | 1.17 | 63.5 | 0.67 | 36.5 |  |  |  |  |
|  | Thaba-Tseka | 4.48 | 57.0 | 1.39 | 17.7 | 2.00 | 25.4 |  |  |
| Residence | Urban | 82.36 | 69.7 | 29.69 | 25.1 | 6.15 | 5.2 | 1.37 | 0.651 |
|  | Rural | 69.54 | 65.6 | 26.15 | 24.7 | 10.27 | 9.7 |  |  |
| Educational level | No education | 5.61 | 36.2 | 7.00 | 45.2 | 2.88 | 18.6 | 12.44 | 0.275 |
|  | Primary | 55.54 | 62.0 | 24.60 | 27.5 | 9.45 | 10.5 |  |  |
|  | Secondary | 80.43 | 77.1 | 19.74 | 18.9 | 4.09 | 3.9 |  |  |
|  | Higher | 10.32 | 69.7 | 4.49 | 30.3 |  |  |  |  |
| Religion | Roman Catholic | 69.50 | 73.9 | 18.39 | 19.6 | 6.14 | 6.5 | 9.36 | 0.321 |
|  | Protestant | 36.93 | 57.7 | 24.00 | 37.5 | 3.06 | 4.8 |  |  |
|  | Pentecostal/other Christians | 37.81 | 70.3 | 9.21 | 17.1 | 6.80 | 12.6 |  |  |
|  | Other religions | 7.66 | 62.1 | 4.24 | 34.4 | 0.43 | 3.5 |  |  |
| Ethnicity | Basotho | 147.34 | 68.6 | 51.61 | 24.0 | 15.77 | 7.3 | 1.72 | 0.489 |
|  | Others | 4.56 | 48.3 | 4.22 | 44.8 | 0.65 | 6.9 |  |  |
| Sex of household head | Male | 103.96 | 61.2 | 50.12 | 29.5 | 15.77 | 9.3 | 11.48 | 0.008* |
|  | Female | 47.94 | 88.3 | 5.72 | 10.5 | 0.65 | 1.2 |  |  |
| Household size | 5 or <5 | 109.02 | 66.9 | 39.83 | 24.4 | 14.18 | 8.7 | 1.36 | 0.575 |
|  | >5 | 42.88 | 70.1 | 16.01 | 26.2 | 2.24 | 3.7 |  |  |
| Media exposure | No media exposure | 12.88 | 40.6 | 14.77 | 46.5 | 4.11 | 12.9 | 22.11 | 0.004* |
|  | Low media exposure | 73.25 | 62.4 | 32.37 | 27.6 | 11.81 | 10.1 |  |  |
|  | High media exposure | 65.78 | 87.7 | 8.70 | 11.6 | 0.51 | 0.7 |  |  |
| Owns a mobile telephone | No | 25.78 | 55.3 | 18.18 | 39.0 | 2.67 | 5.7 | 5.09 | 0.140 |
|  | Yes | 126.12 | 71.0 | 37.66 | 21.2 | 13.75 | 7.7 |  |  |
| Wealth index for urban/rural | Poorest | 22.18 | 56.9 | 15.48 | 39.7 | 1.34 | 3.4 | 11.53 | 0.382 |
|  | Poorer | 28.61 | 66.5 | 10.41 | 24.2 | 3.98 | 9.3 |  |  |
|  | Middle | 34.56 | 65.3 | 10.87 | 20.5 | 7.51 | 14.2 |  |  |
|  | Richer | 19.89 | 63.5 | 9.22 | 29.5 | 2.19 | 7.0 |  |  |
|  | Richest | 46.67 | 80.6 | 9.85 | 17.0 | 1.39 | 2.4 |  |  |
| Occupation | Not working | 31.74 | 74.8 | 6.77 | 16.0 | 3.90 | 9.2 | 16.31 | 0.127 |
|  | Professional/Technical/Managerial | 5.85 | 78.7 | 1.58 | 21.3 |  |  |  |  |
|  | Clerical/Sales/Services | 36.70 | 87.5 | 5.26 | 12.5 |  |  |  |  |
|  | Agricultural workers | 8.23 | 40.0 | 8.88 | 43.2 | 3.45 | 16.8 |  |  |
|  | Manual workers | 69.38 | 62.1 | 33.35 | 29.8 | 9.08 | 8.1 |  |  |
| Currently working | No | 53.41 | 72.7 | 14.04 | 19.1 | 6.00 | 8.2 | 1.61 | 0.570 |
|  | Yes | 98.49 | 65.4 | 41.80 | 27.7 | 10.42 | 6.9 |  |  |
| Total Children | No child | 23.88 | 61.8 | 8.15 | 21.1 | 6.60 | 17.1 | 10.20 | 0.461 |
|  | One child | 36.87 | 71.4 | 11.97 | 23.2 | 2.80 | 5.4 |  |  |
|  | 2-4 children | 75.62 | 73.2 | 22.87 | 22.1 | 4.87 | 4.7 |  |  |
|  | >4 children | 15.53 | 50.9 | 12.84 | 42.1 | 2.16 | 7.1 |  |  |
| Covered by health insurance | No | 124.94 | 68.3 | 43.02 | 23.5 | 15.05 | 8.2 | 1.57 | 0.652 |
|  | Yes | 26.96 |  | 12.82 |  | 1.37 |  |  |  |
| Alcohol use | Never/No | 60.28 | 93.5 | 2.04 | 3.2 | 2.16 | 3.3 | 22.85 | <0.001* |
|  | Yes | 91.61 | 57.4 | 53.80 | 33.7 | 14.26 | 8.9 |  |  |
| Wife beating | No | 125.58 | 72.3 | 39.15 | 22.5 | 8.90 | 5.1 | 7.49 | 0.075 |
|  | Yes | 26.32 | 52.1 | 16.68 | 33.0 | 7.52 | 14.9 |  |  |
| TB-related Stigma | No stigma | 128.27 | 66.5 | 51.42 | 26.7 | 13.16 | 6.8 | 2.04 | 0.517 |
|  | Stigma | 23.63 | 75. | 4.42 | 14.1 | 3.27 | 10.4 |  |  |
| Mental health score | No depression | 114.03 | 71.5 | 37.29 | 23.4 | 8.19 | 5.1 | 12.92 | 0.026* |
|  | Mild | 25.22 | 49.0 | 18.55 | 36.0 | 7.69 | 14.9 |  |  |
|  | Moderate/severe | 12.64 | 95.8 |  |  | 0.55 | 4.2 |  |  |
| Total |  | 151.90 | 67.8 | 55.84 | 24.9 | 16.42 | 7.3 |  |  |

### Predictors of smoking prevalence among people with TB

Being male (AOR=120.15, 95%CI:30.53-472.81, p<0.001), no media exposure (AOR=21.96, 95%CI:5.79-83.31, p<0.001), low media exposure (AOR=4.76, 95%CI:1.33-17.09, p=0.017), alcohol use (AOR=12.64, 95%CI:3.24-49.35, p<0.001), and mild depression (AOR=3.28, 95%CI: 3.28, 95%CI: 1.04-10.38, p=0.043) increased the odds of smoking among TB patients (Table 4). Nevertheless, moderate/severe depression (AOR=0.06, 95%CI: 0.00-0.73, p=0.027) reduced the likelihood of smoking among TB patients.

**Table 4.** Predictors of cigarette smoking among people with TB in Lesotho, 2023-24.

| Parameters |  | COR | 95%<br>Confidence<br>Interval for<br>COR |  | Sig. | AOR | 95%<br>Confidence<br>Interval for<br>AOR |  | Sig. |
| --- | --- | --- | --- | --- | --- | --- | --- | --- | --- |
|  | (Intercept) | 0.01 | 0.00 | 0.06 | <0.001* | 0.00 | 0.00 | 0.02 | <0.001* |
| Gender | Male | 89.25 | 17.30 | 460.36 | <0.001* | 120.15 | 30.53 | 472.81 | <0.001* |
|  | Female | 1.00 |  |  |  | 1.00 |  |  |  |
| Sex of household head | Male | 2.32 | 0.33 | 16.31 | 0.393 |  |  |  |  |
|  | Female | 1.00 |  |  |  |  |  |  |  |
| Media exposure | No media exposure | 20.77 | 4.71 | 91.56 | <0.001* | 21.96 | 5.79 | 83.31 | <0.001* |
|  | Low | 4.24 | 1.19 | 15.10 | 0.026* | 4.76 | 1.33 | 17.09 | 0.017* |
|  | High | 1.00 |  |  |  | 1.00 |  |  |  |
| Occupation | Not working | 0.78 | 0.20 | 3.02 | 0.720 |  |  |  |  |
|  | Professional/Technical/Managerial | 1.11 | 0.18 | 7.01 | 0.908 |  |  |  |  |
|  | Clerical/Sales/Services | 0.41 | 0.06 | 2.68 | 0.347 |  |  |  |  |
|  | Agricultural workers | 3.84 | 0.80 | 18.35 | 0.091 |  |  |  |  |
|  | Manual workers | 1.00 |  |  |  |  |  |  |  |
| Alcohol use | Yes | 18.11 | 3.16 | 103.75 | 0.001* | 12.64 | 3.24 | 49.35 | <0.001* |
|  | No | 1.00 |  |  |  | 1.00 |  |  |  |
| Wife beating | No | 0.36 | 0.12 | 1.11 | 0.074 |  |  |  |  |
|  | Yes | 1.00 |  |  |  |  |  |  |  |
| Mental health status | Moderate/severe | 0.02 | 0.00 | 0.18 | <0.001* | 0.06 | 0.00 | 0.73 | 0.027* |
|  | Mild depression | 2.49 | 0.68 | 9.14 | 0.168 | 3.28 | 1.04 | 10.38 | 0.043* |
|  | No depression | 1.00 |  |  |  | 1.00 |  |  |  |

\*Significant at p-value < 0.05, COR = Crude odd ratio, AOR = Adjusted odd ratio

### Predictors of daily smoking among people with TB

Being male (AOR=234.07, 95%CI:108.69-504.08, p<0.001), no media exposure (AOR=18.93, 95%CI:4.58-78.28, p<0.001), and alcohol use (AOR=21.26, 95%CI:3.79-119.23, p=0.001) increased the odds of smoking among TB patients (Table 5). In contrast, moderate/severe depression (AOR=0.00, 95%CI: 0.00-0.00, p<0.001) reduced the likelihood of smoking among TB patients.

**Table 5.**
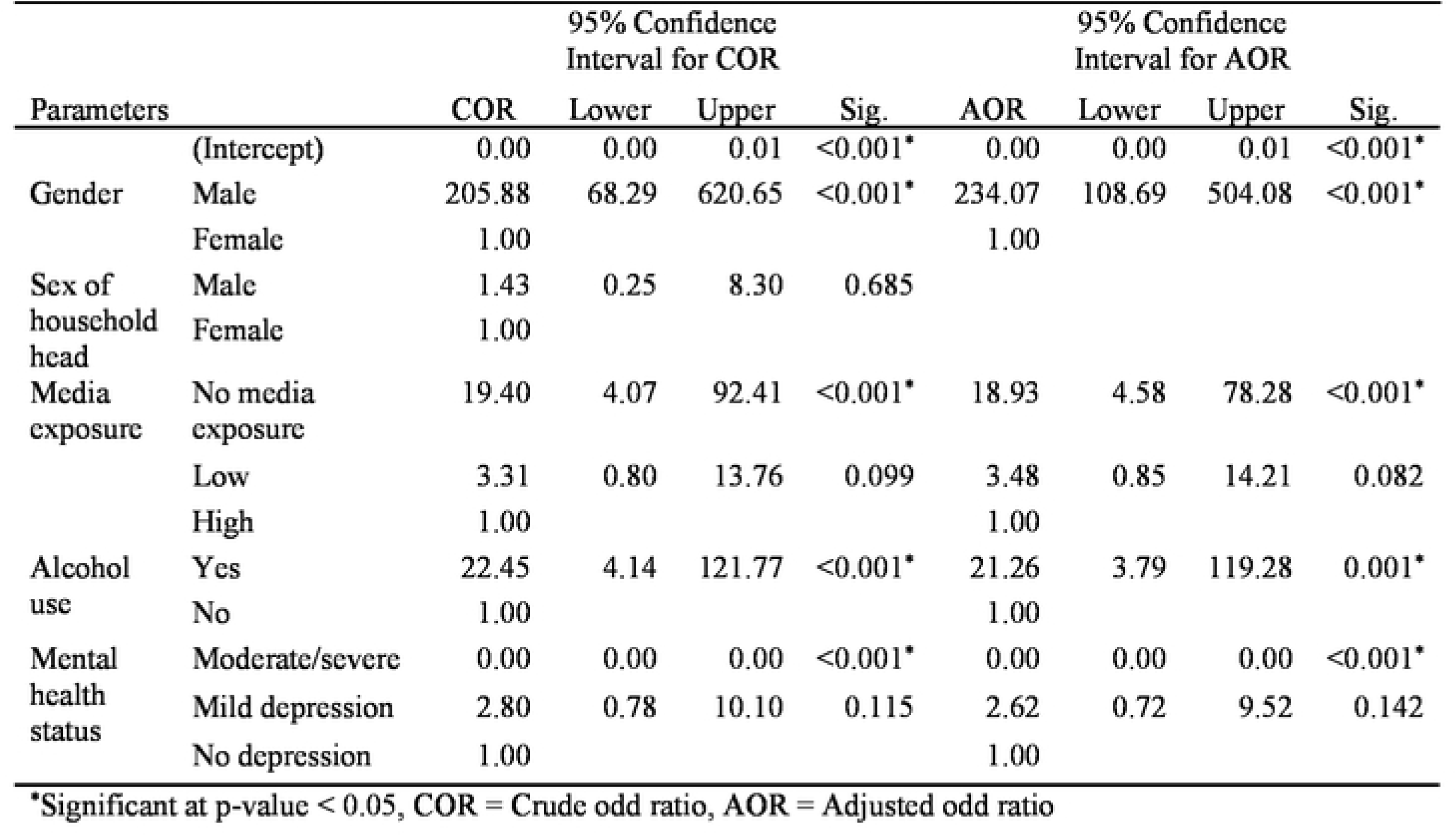
Predictors of daily cigarette smoking among people with TB in Lesotho, 2023-24.

### Predictors of occasional smoking among people with TB

Being male (AOR=46.55, 95%CI:4.69-461.61, p=0.001), no media exposure (AOR=94.66, 95%CI:7.60-1179.67, p=0.001), and low media exposure (AOR=26.34, 95%CI:2.54-272.98, p=0.007), and mild depression (AOR=6.18, 95%CI:1.43-26.66, p=0.015) increased the odds of smoking among TB patients (Table 6).

**Table 6.**
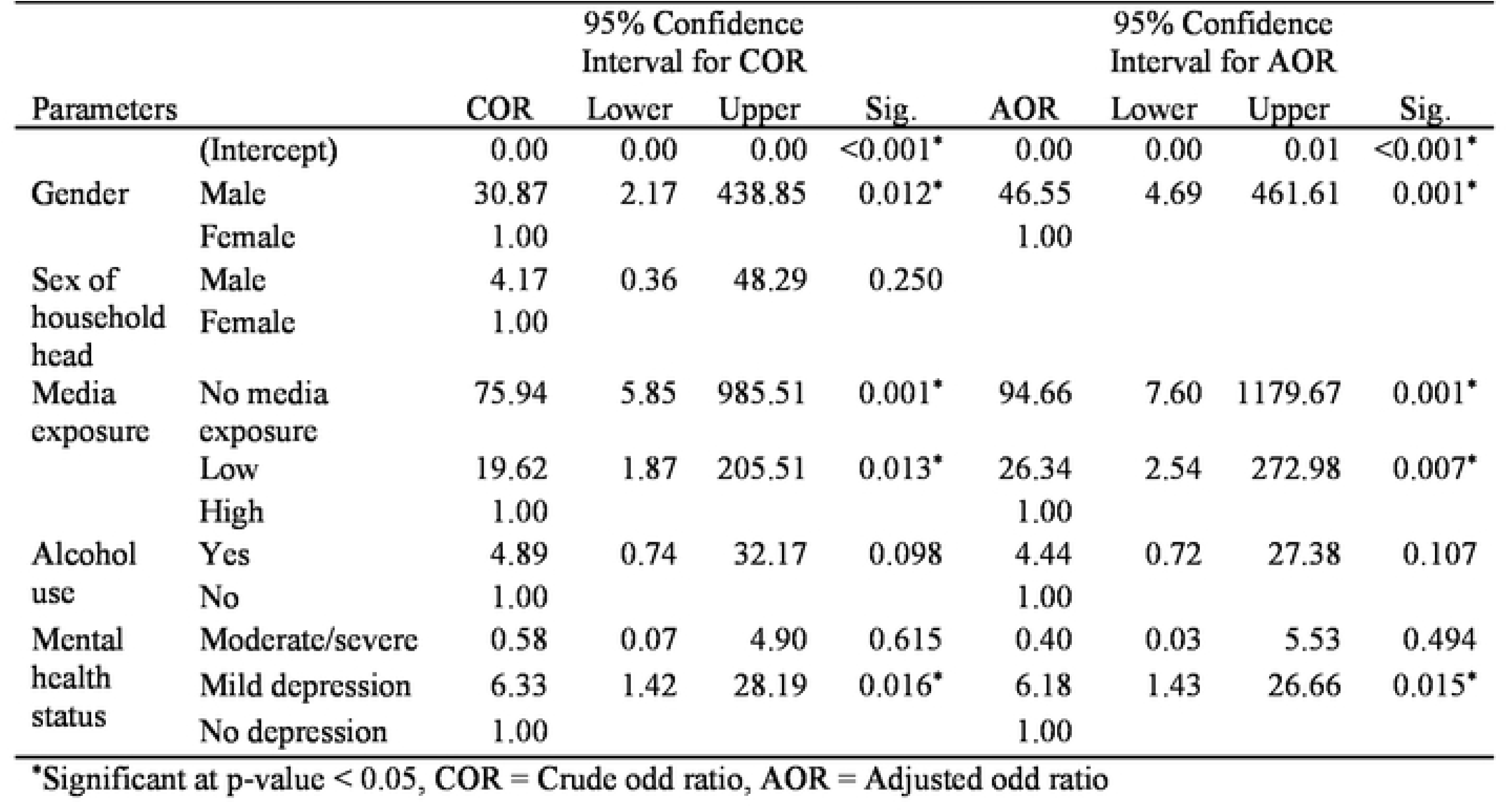
Predictors of occasional cigarette smoking among people with TB in Lesotho, 2023-24.

| Parameters |  | 95% Confidence Interval for COR |  |  | Sig. | 95% Confidence Interval for AOR |  |  | Sig. |
| --- | --- | --- | --- | --- | --- | --- | --- | --- | --- |
|  |  | COR | Lower | Upper |  | AOR | Lower | Upper |  |
|  | (Intercept) | 0.00 | 0.00 | 0.00 | <0.001* | 0.00 | 0.00 | 0.01 | <0.001* |
| Gender | Male | 30.87 | 2.17 | 438.85 | 0.012* | 46.55 | 4.69 | 461.61 | 0.001* |
|  | Female | 1.00 |  |  |  | 1.00 |  |  |  |
| Sex of household head | Male | 4.17 | 0.36 | 48.29 | 0.250 |  |  |  |  |
|  | Female | 1.00 |  |  |  |  |  |  |  |
| Media exposure | No media exposure | 75.94 | 5.85 | 985.51 | 0.001* | 94.66 | 7.60 | 1179.67 | 0.001* |
|  | Low | 19.62 | 1.87 | 205.51 | 0.013* | 26.34 | 2.54 | 272.98 | 0.007* |
|  | High | 1.00 |  |  |  | 1.00 |  |  |  |
| Alcohol use | Yes | 4.89 | 0.74 | 32.17 | 0.098 | 4.44 | 0.72 | 27.38 | 0.107 |
|  | No | 1.00 |  |  |  | 1.00 |  |  |  |
| Mental health status | Moderate/severe | 0.58 | 0.07 | 4.90 | 0.615 | 0.40 | 0.03 | 5.53 | 0.494 |
|  | Mild depression | 6.33 | 1.42 | 28.19 | 0.016* | 6.18 | 1.43 | 26.66 | 0.015* |
|  | No depression | 1.00 |  |  |  | 1.00 |  |  |  |

\*Significant at p-value < 0.05, COR = Crude odd ratio, AOR = Adjusted odd ratio

## Discussion

The purpose of this study was to determine the prevalence and correlates of smoking among people with TB in Lesotho. Our findings that the high prevalence of smoking among people with TB is associated with male gender, media exposure, alcohol consumption and mental health status warrant further exploration.

This study found that almost a third of people with TB smokes, a much higher figure than the 18% smoking prevalence in Lesotho’s general population[3]. Since smoking behaviour and impairs immunity and increased the chances of developing TB[16], the high prevalence of TB patient smokers might explain the increasing number of TB cases attributable to smoking in Lesotho[2]. The smoking prevalence in the current study is consistent with evidence from Bangladesh[24], but higher than the findings from other studies in India, Iran, Pakistan, Uganda and Ethiopia[18, 23, 24, 27, 31]. In contrast, our finding is lower than the prevalence from prior studies from Gabon, Spain, Brazil, Georgia, Bangladesh, Jordan, Uganda, South Africa, and Botswana[7, 10, 17, 19, 24–26, 28–30, 32]. The inter-country differences could be due to baseline differences in the smoking habits of the general population from which the TB patients’ subpopulations were derived.

Our finding of high TB patient smokers might explain the increasing number of TB cases attributable to smoking in Lesotho[2]. The high prevalence of TB patient smokers may be due to limited smoking control measures or a higher addiction to nicotine among people with TB. Notwithstanding signing the World Health Organization’s Framework Convention on Tobacco Control, the Government of Lesotho lacks tobacco packaging regulations and regulations on tobacco advertising, promotion and sponsorship. While a restricting or eliminating marketing is key to tobacco control success, Lesotho’s government has not implemented any component of the direct and indirect bans on TAPS. Similarly, the weak tobacco tax policies incentivize tobacco users by making tobacco products affordable. Additionally, the Government of Lesotho need to integrate smoking cessation counselling into TB care to help people with TB quit smoking.

Consistent with evidence from previous studies[10, 16, 25, 30, 32–34], our study finding revealed a higher likelihood of smoking, daily smoking and occasional smoking among males with TB, In the current study, 57.6% of men with TB were smokers, compared to 39.2 % of males in the general population. Contrastingly, we found that 3.3% of women with TB smoked compared to 0.4% in the general population. The lower prevalence of smoking among women with TB in this study is not unsurprising given the cultural situation of women in most sub-Saharan African settings where strong social norms and taboos which discourage women to smoke and cigarette smoking is deemed an inappropriate behaviour for women, while smoking among men in is acceptable, and represents a symbol of status and social power[34, 45]. The significant association of smoking with male gender might account for the gender differences in TB prevalence with more male TB cases than females[10, 16].

We also found that a lack of or low media exposure predicted smoking among people with TB in Lesotho. While people with TB who lacked media exposure were nine times more likely to smoke, low media exposure resulted in four-fold likelihood of smoking among people with TB. A lack of media exposure also increased the odds of daily and occasional smoking. Our findings imply a beneficial influence of media exposure on reducing smoking prevalence among people with TB. Although no prior study has assessed how exposure to tobacco-related media influences smoking behaviour among people with TB, evidence indicates that high media exposure lowers the risk of cigarette smoking among adults in the general population[46, 47]. Exposure to anti-smoking messages was associated with greater risk perceptions of smoking, which could facilitate prevention of smoking initiation and intention to quit[48]. Nevertheless, tobacco control in Africa, including Lesotho, is constrained by limited use of best-practice anti-tobacco media campaign, the competition to use the media to push for or against tobacco control, a lack of long-term plan for anti-tobacco media, inadequate funding for mass media interventions, and low involvement of target beneficiaries in the process of developing anti-tobacco media products[49, 50].

In this study, alcohol consumption increased the likelihood of smoking and daily smoking among people with TB. Similarly, alcohol predicted a higher odd of smoking among people with TB in previous studies[10, 16, 34, 35]. Our finding mirrors the influence of alcohol use on smoking in the general population reported in other African studies[51, 52]. Alcohol consumption increases the urge and impulse to smoke by disrupting cognitive functions, weakening self-control or self-regulation, interacting with the brain nicotinic receptors and activating dopamine release[53]. In the current study, about 33% of people with TB using alcohol smoked compared to about 6% of people with TB, who do not use alcohol. This evidence will enable the National TB Program in Lesotho to target TB patients, who use alcohol, with smoking cessation interventions.

While mild depression increased the likelihood of smoking and occasional smoking, moderate to severe depression decreased the odds of smoking and daily smoking among people with TB. The influence of mild depression on smoking status in our finding is consistent with prior studies showing that depressive symptoms increased the risk of smoking among TB patients[32]. Long TB treatment period can lead to mental and emotional problems, necessitating smoking. Smokers, including people with TB, use nicotine to medicate their depressed mood because of the reinforcing effects of nicotine’s mood-altering characteristics[54]. Nicotine, the main content of tobacco, causes the release of dopamine, which improves mood and reduces mental stress[32, 54]. Our finding that moderate to severe depression decreased the odds of smoking and daily smoking among people with TB was surprising because moderate to severe depression deceased the likelihood of smoking cessation[55]. Nevertheless, the significant decrease in the odds of smoking in moderate to severely depressed TB patients in our study may be due to increased awareness of the negative impact of smoking on TB treatment outcome, improved access to smoking cessation during TB treatment, and knowing that quitting smoking can improve mental well-being[35, 56].

The main strength of this study was using a nationally representative, population-level data to estimate the prevalence and determinants of smoking among people with TB, increasing the likelihood of generalizing our findings. Besides social characteristics of respondents, our study explored the influence of TB-related stigma and mental health status of people with TB on smoking status. Nonetheless, being a cross-sectional survey, the study can only show association between smoking and the determinants and cannot establish causality.

## Conclusion

The purpose of this study was to determine the prevalence of smoking among people with TB and its associated factors in Lesotho. The prevalence of smoking among people with TB is high in Lesotho. Male gender is a risk factor for smoking among people with TB and might account for the gender differences in TB prevalence in Lesotho. Limited media exposure increases the risk of smoking, highlighting a need for long-term plan for best-practice anti-tobacco media campaign to push for tobacco control, and adequate funding for mass media interventions. Alcohol use predisposed to smoking among people with TB, underscoring the intersection of alcohol use and smoking in the social epidemiology of TB. While mild depression increased the odds of cigarette smoking and occasional smoking among people with TB, moderate/severe depression reduced the likelihood of smoking and daily smoking among TB patients. Policymakers and health system practitioners should consider the identified factors for implementing effective interventions to reduce smoking initiation and increase smoking cessation among people with TB in Lesotho.

## Data Availability

The data underlying the results presented in the study are available from https://dhsprogram.com/data/available-datasets.cfm

